# By the numbers and in their own words: A mixed methods study of unmet needs and humanitarian inclusion of older Syrian refugees in Lebanon

**DOI:** 10.1101/2024.04.02.24305052

**Authors:** Sarah Al Omari, Gladys Honein-AbouHaidar, Abla Mehio Sibai

## Abstract

Older people constitute an overlooked vulnerable population in humanitarian crises. Lebanon is a small country that hosts the largest number of refugees per capita in the world. With exacerbating socioeconomic conditions, exclusionary policies against refugees, and a fragmented humanitarian system, the status of older Syrian refugees (OSRs) requires special attention. This study aimed to explore OSRs’ unmet needs, coping strategies, available humanitarian services, and some indicators of the humanitarian inclusion standards focusing on the shelter, health, nutrition and food security, and water, sanitation and hygiene sectors. We conducted a convergent mixed-methods study between December 2021 and March 2022 in the North and Bekaa, including a cross-sectional survey with 461 participants and 14 semi-structured interviews. Results show that OSRs lived in inappropriate shelters (cold, leaking rainwater), especially in informal tented settlements. High rent prices pushed refugee households with elderly to prioritize paying rent at the expense of other needs such as food and medication, particularly when food cash transfer is the sole source of income, jeopardizing food security and intake. Access to dignifying and accessible bathing facilities was compromised in ITSs with shared facilities. Substantial medical costs hindered OSRs access to healthcare such as surgeries. Due to the crisis, chronic medications are not always available in dispensaries for subsidized cost, pushing OSRs to non-compliance and selling food assistance to buy medications. Soaring fuel prices hindered OSRs access to heating and transportation to receive healthcare. No efforts were reported in collecting data on OSRs’ needs, targeting them with information on services, or soliciting feedback for programming, especially in the absence of any age-tailored interventions. Findings shed light on the precarious living conditions of OSRs in Lebanon and add to the body of evidence documenting their invisibility to the humanitarian response. An age-inclusive response is needed through holistic, tailored, and sustainable interventions.

## Introduction

The intersection of accelerating population ageing with steady rise in man-made and natural disasters poses tremendous risks on older populations in low- and middle-income countries disproportionately affected by disasters [1–3]. Older populations in disaster settings are particularly vulnerable, making up to 4% of UNHCR’s overall population of concern [4]. The intersection between the normative challenges of ageing, such as the significant decline in physical, cognitive, and psychosocial wellbeing on one hand, and the horrific circumstances of disasters and forced migration on the other hand, ranks elderly populations as a highly vulnerable segment with very specific needs. Risks that older people face in disasters include being left behind, physical injury, mental traumatization and illness, loss of loved ones either through death or separation, loneliness, and the loss of economic opportunities, social status and roles [5–7].

A multitude of policies and guidelines developed by international organizations acknowledge the particularities of elderly’s needs in crises and the approaches to ensure an inclusive response at all levels of programming [6, 8–10]. Unfortunately, the gap between policies and action is too wide, showing a systematic exclusion of elderly refugees from the humanitarian response, a phenomenon that has been attributed to ageist perceptions and practices in the humanitarian realm [5, 6, 11–15]. Exclusion is marked at different stages of the response. Specific older-age policies are lacking in many organizations, data collection is not disaggregated by age at pre-assessment levels, programming is not age-conscious especially in health and nutrition, and monitoring and evaluation activities do not report on indicators specific to elderly, or present age-disaggregated findings to assess program performance [12, 15]. At another level, the proportion of humanitarian funding channeled towards the needs of elderly refugees for elderly refugees is negligible [16].

The situation becomes grimmer when nested within a multi-dimensional crisis, as is the case in Lebanon. Lebanon, this small middle income country bordering Syria, hosts the largest number of refugees per capita in the world, with an estimated 1.5 million Syrian refugees, of whom 2.6% are above 60 years of age [17]. Lebanon is not signatory on the 1951 Refugee Convention, and its refugee legal policies and legislations are exclusionary [18–21]. Since September 2019, Lebanon has been suffering from a myriad of overlapping crises, including dramatic currency devaluation, collapse of many public services, political turmoil, the COVID-19 pandemic effects, and the infamous Beirut Blast, all exacerbating a protracted refugee crisis that has strained pre-existing fragmented systems.

In order to strengthen the accountability of humanitarian actors towards older people and people with disabilities within a clearly defined framework, the Age and Disability Consortium as part of the Age and Disability Capacity Program (ADCAP) published in 2018 the “Humanitarian Inclusion Standards for Older People and People with Disabilities” [22]. These standards (HISs) aim to guide humanitarian action in responding to the needs and leveraging the capacities of older people and people with disabilities, ensuring an inclusive and participatory humanitarian response, from organizational policies and advocacy to programming and field training. The nine inclusion standards include: identification, safe and equitable access, resilience, knowledge and participation, feedback and complaints, coordination, learning, human resources, and resources management. In addition, a set of seven sector-specific standards are presented for shelter, settlement and household items, health, education, nutrition, food security and livelihoods, and protection. These sector-specific standards are mainly designed around three areas, which can be mapped under some of the nine standards: data and information management (under identification), addressing barriers (under safe and equitable access), and participation of older people and people with disabilities and strengthening of their capacities (under knowledge and participation, and resilience). In 2020, it was made part of the Sphere Humanitarian Standards Partnership [22].

While multiple studies have been conducted with older Syrian refugees (OSRs) in Lebanon [14, 23–26], and other neighboring countries [27–30], an extensive needs assessment with a focus on their reports and perceptions of inclusion is missing. This study guided by a human-rights lens is looking at a population with a myriad of intersecting vulnerabilities to emphasize its right to access humanitarian assistance. It aims to inform about the existing unmet needs and the status of humanitarian inclusion of OSRs in Lebanon. A convergent parallel mixed methods design is used, where both quantitative and qualitative data are collected in parallel, analyzed separately, and then merged [31].

The quantitative component in this study will measure the distribution of unmet needs, as well as indicators of humanitarian assistance and inclusion across four sectors: shelter, health, nutrition, and water, sanitation, and hygiene (WASH). The qualitative component will explore OSRs perceptions and experiences with unmet needs and their assessment of access to assistance. The reason for collecting both types of data is to validate and triangulate findings and to bring greater insight than would be obtained by either type exclusively.

In other words, we aim in this study to answer the following questions:

What are the unmet needs of OSRs in Lebanon in the shelter, health, nutrition, and WASH sectors? What are existing coping strategies given the existing adversities?
What types of assistance are OSRs receiving in these sectors? What assistance can’t they access? Why?
To what extent is the humanitarian response across the different sectors inclusive of OSRs as reflected through a number of indicators constructed from the ADCAP humanitarian inclusion standards?

## Methods

A convergent parallel mixed methods design with equally weighted quantitative and qualitative strands was used. Both quantitative and qualitative data were collected in parallel, analyzed separately, and then merged [31]. In both arms of the study, two main themes were explored, namely unmet needs and inclusion in humanitarian services and assistance. While a set of detailed indicators and questions were used to inquire about issues and unmet needs in both arms, the HISs were used as a guiding framework to measuring and characterizing the inclusion status of OSRs on the humanitarian response in Lebanon. For the quantitative arm, a set of indicators were constructed under each of the HIS’s: identification, safe and equitable access, knowledge and participation, feedback and complaints, and learning. Some of the formulated indicators were general, while others were sector-specific under shelter, health, nutrition, food security, or WASH. The same topics were also inquired and probed about in the qualitative interviews, soliciting views, perceptions, and experiences. The content of the research tools is detailed below.

### Setting of the study

The study was carried out in December 2021 and March 2022 in Akkar, North, and Bekaa governorates in Lebanon. Those three areas contain an estimated 527,000 refugees, which is approximately 66% of the total Syrian refugee population in Lebanon [32]. Because the Lebanese government prevents the establishment of official refugee camps, Syrian refugees reside either in informal tented settlements often established on private, agricultural land, or rent private accommodations in towns and cities [19]. Almost 22%, 45%, and 8% of the population lives in informal tented settlements (ITSs), and 62%, 50% and 75% live in hosted communities in each of Akkar, Bekaa, and North respectively [33].

### Quantitative component

#### Sampling and recruitment

A convenient and purposeful non-probability sampling strategy has been employed due to the lack of a sampling frame. We recruited a local humanitarian agency active in providing field services in the Akkar, Bekaa, and North governorates to allow us access to the refugee communities and collect data. At a first stage, the agency was instructed by the research team to provide a list of recruitment sites, both informal tented settlements and hosted community clusters, that are also diverse in terms of humanitarian service coverage and living conditions. The list also included an enumeration of older Syrian refugees aged 50 or above per site through the agency’s focal points. The final total sample size of 461 was determined by budget and time constraints.

#### Survey tool

The survey instrument consisted of several sections. It began with sociodemographic, socioeconomic, and household information. Next, we included a section on humanitarian assistance including questions on data collection, data access, and received assistance. Furthermore, sections for each of the sectors of inquiry were included: shelter, health, nutrition and food security, and WASH. Under each of these sections, we questions inquired about unmet needs, as well as sector-specific inclusion indicator questions.

Under health, the following constructs were solicited to assess needs: experiencing pain, disability including activities of daily living (ADL) and instrumental activities of daily living (IADL), chronic diseases, adherence to medications, and associated barriers. We incorporated and adapted questions from the Katz index for ADL, the Lawton index for IADL, and the Washington Group scale for disability to assess disability [34–36]. Finally, we inquired about access to primary healthcare (PHC) and hospitalization.

As for nutrition and food insecurity, indicators from the HelpAge’s Rapid Assessment Method for Older People were used including: meal frequency, food diversity and nutrient consumption, and food coping strategies [37]. Indicators under food group consumption were compiled to form the Diet Diversity Score, a measure that ranges from 0 to 11 and consists of the number of food groups consumed in the last 24 hours. Furthermore, the Household Hunger Scale score, a measure of severe food insecurity, was calculated based on question on food coping strategies. The scale categorizes the household in which the elderly lives into living under either “extreme hunger”, “moderate hunger”, and “little or no hunger” [37]. Finally, under shelter and WASH, OSRs were asked for their assessment of any problems with the shelter, water, latrines, as well as any humanitarian assistance and barriers.

The survey tool was uploaded into KOBO toolbox and data was collected on electronic tablets.

#### Data collection

A schedule for visiting the closer, then farther sites was set up with the research team, and the participants were purposefully approached such that the sample would produce an equal distribution by gender, a wide age range, and include home-bound older refugees. Two teams of four experienced data collectors each were mobilized to cover Akkar and North on one hand, and Bekaa on the other, and worked in parallel. All enumerators underwent a 2-day training workshop. Data collection began on December 16^th^ and ended on the 31^st^. The survey consisted of face-to-face structured interviews; each interview took approximately 30-40 minutes. All field activities were supervised by members of the research team. In addition, the data collection procedure and tool were pilot-tested for two full days in both Akkar and Bekaa areas simultaneously, completing 36 interviews. This led to another round of training and tool refinement. Surveys collected in the pilot phase were not included in the analyses.

#### Data analysis

Descriptive statistics: Continuous variables were summarized using means and standard deviations, while proportions were presented to describe categorical variables.

Inferential statistics: Bivariate analysis employed Student’s t-test to explore associations between a binary predictor and a continuous outcome. Chi-square test, Fisher’s exact test, and Cochrane Armitage trend tests were employed to test for relations between categorical variables. Binary and ordinal logistic regressions were performed to test for gender differences in binary and categorical outcomes while controlling for age. All tests were reported at the 0.05 significance level. Data were manipulated using Stata version 17.

### Qualitative component

A descriptive qualitative methodology was employed to explore participants’ perceptions and views on unmet needs and inclusion in humanitarian assistance and triangulate them with findings from the survey [38, 39].

#### Sampling and Recruitment

We purposefully selected a subsample of 14 informative participants from the quantitative component. The sample was also purposefully recruited to have a balanced gender composition and to include participants from both ITSs and hosted communities in the North and Bekaa areas wherever convenient.

#### Topic guide

A topic guide of semi-structured nature was prepared and included questions on: living conditions and unmet needs across the sectors of interest, coping strategies employed by the elderly, and inquiring about received assistance and humanitarian services offered to older refugees. Participants were probed to provide details on the difficulties they face accessing basic needs, facilitators and barriers towards accessing humanitarian assistance, and whether the humanitarian response is tailored to the needs of elderly at the data collection, programming, and accountability levels. In other words, the same HISs covered in the quantitative indicators were explored qualitatively in the semi-structured interviews, namely identification, safe and equitable access, knowledge and participation, feedback and complaints, and learning.

#### Data collection

Data were collected through audio-recording the face-to-face semi-structured interviews with the sub-sample of 14 participants between the 2^nd^ and 29^th^ of March, 2022. In-depth interviews were carried out sequentially and stopped when data saturation was reached. Data was collected by the first author who conceptualized and designed the tools, and notes were taken during interviews by a trained and experienced research assistant. Both data collectors speak Arabic as their mother tongue and can fully communicate in the local Syrian dialect used by the OSRs. Interviews took place in the OSRs’ households and after ensuring that only the elderly and the research team were present in the room.

#### Data analysis

Qualitative interviews recordings were first transcribed in verbatim, then transcriptions were analyzed using thematic analysis. Transcripts were coded deductively under concepts of unmet needs, coping strategies, and access to humanitarian assistance and services. Codes were then combined into sub-themes and themes to answer the research questions. Under each sector of interest, codes on existing issues, unmet needs, coping strategies, access to assistance, and inclusion concepts were extracted and analyzed.

### Integration and reporting of quantitative and qualitative arms

We used the *merging* approach to integrate the results of the quantitative and qualitative components after they were both analyzed separately. Mixed methods results are reported in using a narrative *weaving* approach, where quantitative and qualitative results are written together on a theme-by-theme basis [31, 40]. Integration was conducted manually by the primary author (SAO) and validated by the second author (GHA). Findings were further discussed for concordance, discordance or expansion. Findings were reported based on Good Reporting of A Mixed Methods Study (GRAMMS) criteria (S1 Appendix) [41].

### Ethical considerations and clearance

The study was approved by the Social and Behavioral Institutional Review Board at the American University of Beirut (ID: SBS-2021-0214). Due to COVID-19 restrictions at the time of data collection, verbal informed consent was solicited from each of the participants in both the quantitative and the qualitative components as dictated by the Board.

## Results

### Sample Profile: Sociodemographic and Household Information

All sociodemographic characteristics of the survey sample are summarized in Table 1. A total of 461 OSRs participated in the survey with a median age of 57 years (IQR=11 years), with 59.65% from Bekaa, 29% from the North, and 40.35% from Akkar. The majority (73%) resided in ITSs, were female, both overall (56%) and per governorate, and most participants were married (70.5%) and illiterate (55.1%). Only 11.5% reported working, with significantly higher proportion of working males than females (p<0.001). 96.7% belonged to households having an income of less than 75 USD per month, and 87% reported being in debt. Only 40 participants (8.7%) reported living alone in the household, out of those the majority were women (67.5%). Excluding households of elderly living alone, 86.5% of households were elderly headed, 70.6% of which being male-headed. Female heads of households (25.4%) were predominantly married (41%) or widowed (41%). Almost 55% of the households reported an elderly as the main breadwinner, 14% of which were female elderly.

**Table 1:** Summary sociodemographic and household characteristics of the survey sample.

|  | n | % |
| --- | --- | --- |
| Age (median (IQR)) |  |  |
| Area |  |  |
| North | 186 | 40.35% |
| Bekaa | 275 | 59.65% |
| Governorate |  |  |
| Akkar | 52 | 11.28% |
| North | 134 | 29.07% |
| Bekaa | 275 | 59.65% |
| Site type |  |  |
| ITS | 336 | 72.89% |
| Hosted | 125 | 27.11% |
| Gender |  |  |
| Male | 203 | 44.03% |
| Female | 258 | 55.97% |
| Education level |  |  |
| Illiterate | 254 | 55.10% |
| Reads and writes | 49 | 10.63% |
| Elementary | 77 | 16.70% |
| Middle school | 53 | 11.50% |
| High school | 24 | 5.21% |
| College/university | 4 | 0.87% |
| Marital status |  |  |
| Never married | 11 | 2.39% |
| Married | 325 | 70.50% |
| Widowed | 97 | 21.04% |
| Divorced | 9 | 1.95% |
| Separated | 19 | 4.12% |
| Type of elderly-headed household |  |  |
| Male-elderly-headed household | 270 | 58.57% |
| Female-elderly-headed household | 134 | 29.07% |
| None-elderly-headed-household | 57 | 12.36% |
| Elderly is main breadwinner in the household | 180 | 39.22% |
| The elderly has a caregiving role | 263 | 62.47% |
| Elderly lives alone | 40 | 8.68% |
| Elderly works | 53 | 11.50% |
| Household income |  |  |
| <500,000 LBP (<18.5 USD) | 92 | 20.26% |
| 500,000 - 1,000,000 LBP (18.5-37 USD) | 100 | 22.03% |
| 1,000,000 - 1,500,000 LBP (37-55.56 USD) | 121 | 26.65% |
| 1,500,000 - 2,000,000 LBP (55.56-74 USD) | 75 | 16.52% |
| >2,000,000 LBP (>74 USD) | 58 | 12.78% |
| Don't know | 8 | 1.76% |
| Presence of household debt | 403 | 87.42% |
| Debt amount |  |  |
| <500,000 LBP (<18.5 USD) | 14 | 3.47% |
| 500,000 - 1,000,000 LBP (18.5-37 USD) | 33 | 8.19% |
| 1,000,000 - 1,500,000 LBP (37-55.56 USD) | 32 | 7.94% |
| 1,500,000 - 2,000,000 LBP (55.56-74 USD) | 38 | 9.43% |
| >2,000,000 LBP (>74 USD) | 280 | 69.48% |
| Don't know | 6 | 1.49% |
| Primary source of income |  |  |
| Transfers from relatives in Lebanon or abroad | 6 | 1.30% |
| Work wage | 158 | 34.27% |
| Humanitarian assistance | 234 | 50.76% |
| Begging | 3 | 0.65% |
| Savings | 1 | 0.22% |
| Debt | 19 | 4.12% |
| Selling assets | 3 | 0.65% |
| Selling assistance | 2 | 0.43% |
| Assistance from charity | 27 | 5.86% |
| Other | 6 | 1.30% |
| Don't know | 2 | 0.43% |
| Secondary source of income |  |  |
| Transfers from relatives in Lebanon or abroad | 4 | 0.87% |
| Work wage | 77 | 16.70% |
| Humanitarian assistance | 157 | 34.06% |
| Begging | 2 | 0.43% |
| Savings | 9 | 1.95% |
| Debt | 146 | 31.67% |
| Selling assets | 2 | 0.43% |
| Selling assistance | 6 | 1.30% |
| Assistance from charity | 41 | 8.89% |
| Other | 5 | 1.08% |
| Don't know | 12 | 2.60% |

A total of 14 participants were consulted in the qualitative component. Their ages ranged between 53 and 82 years old, and 9 (64.3%) were woman. The majority live in ITSs (n=10, 71.4%). Table 2 shows a description of the main characteristics of the qualitative component participants.

**Table 2:**
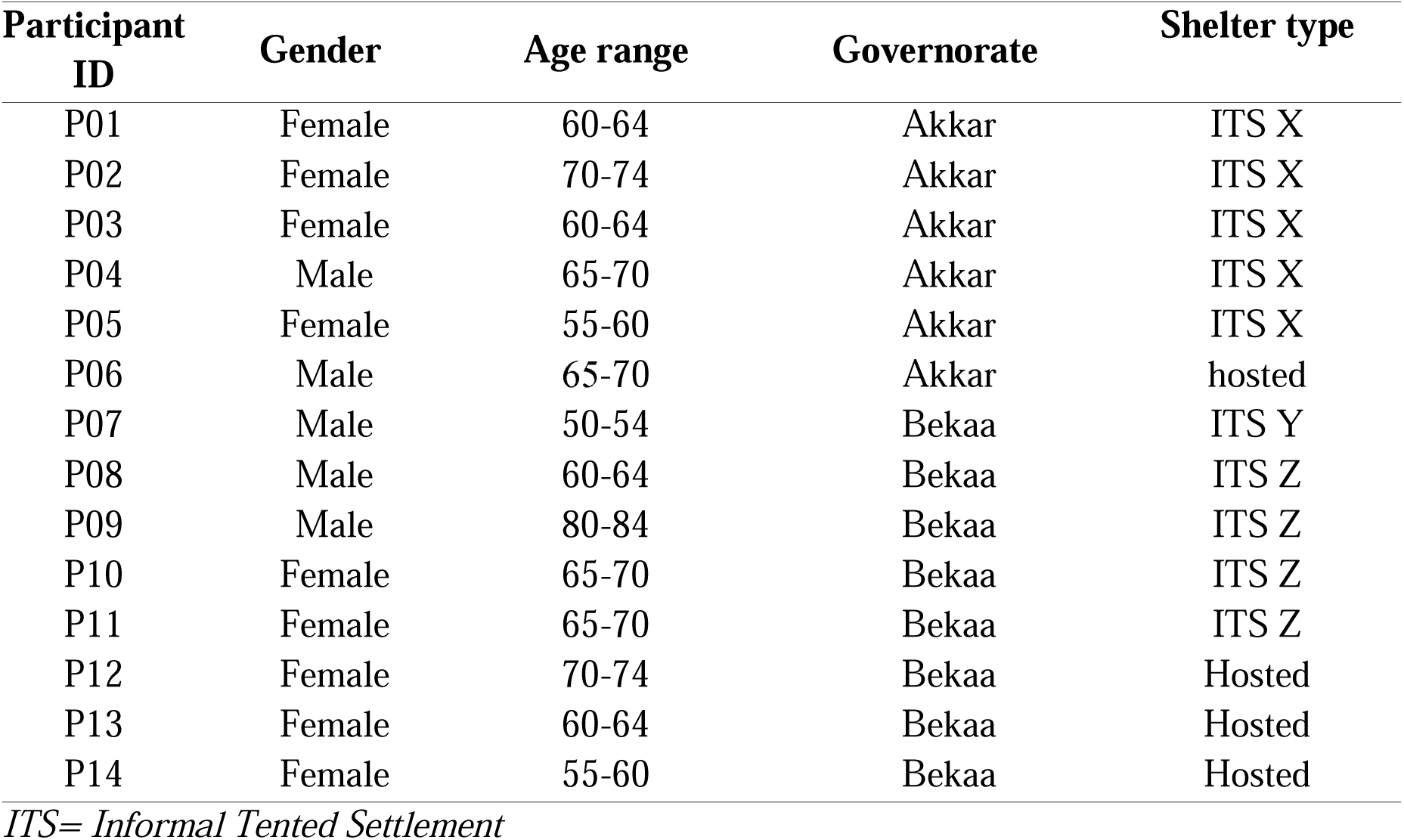
Characteristics of participants in the qualitative interviews.

### General Feedback on Humanitarian Assistance

Table 3 summarized survey participants’ responses on receipt and general assessment of humanitarian assistance.

**Table 3:**
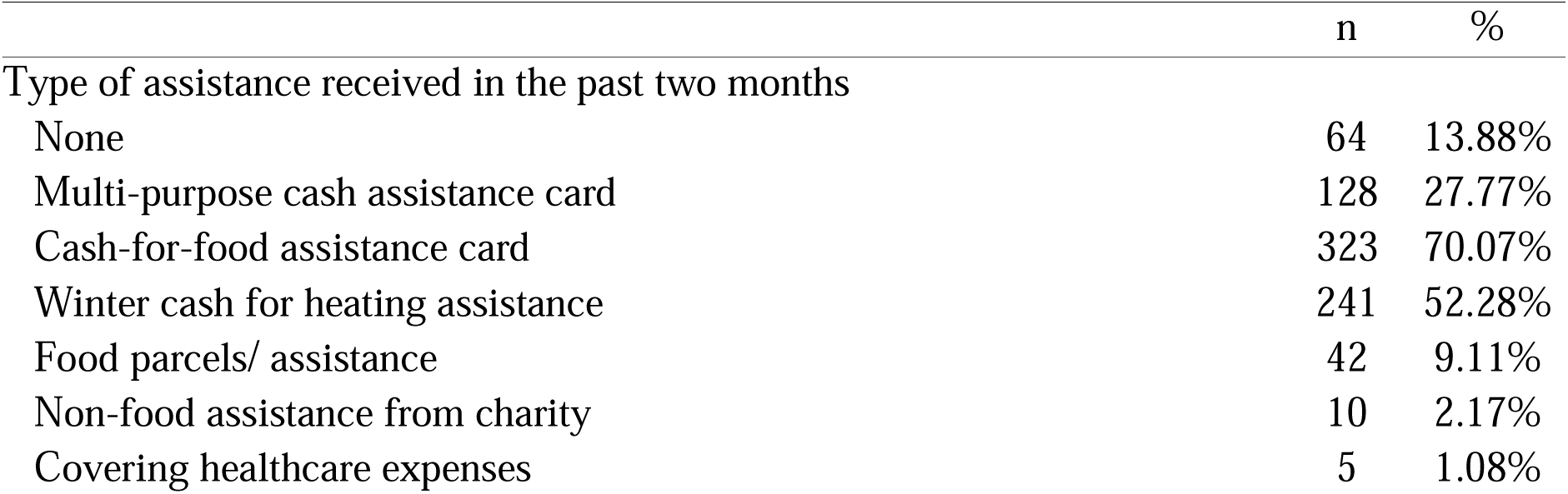
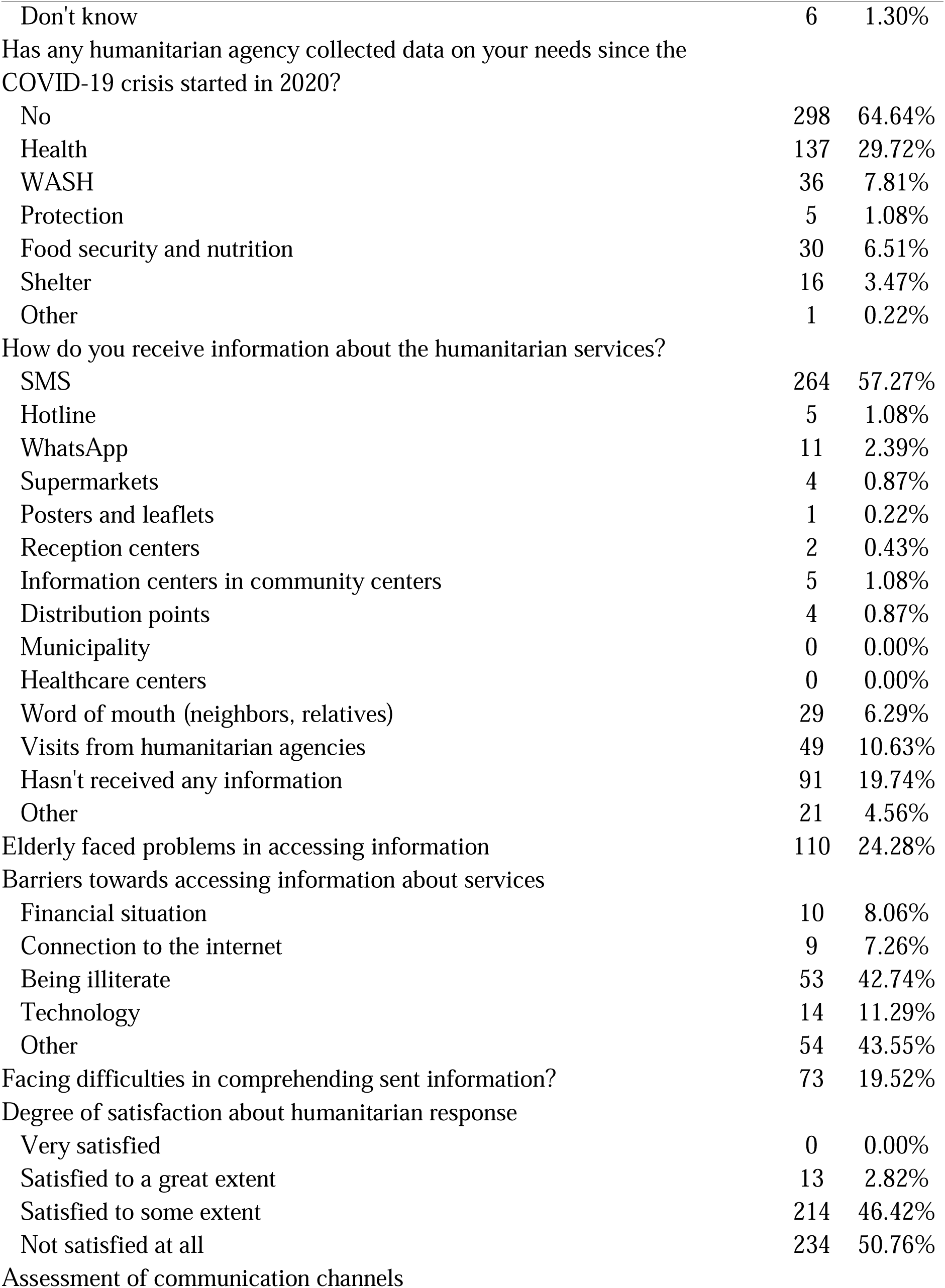
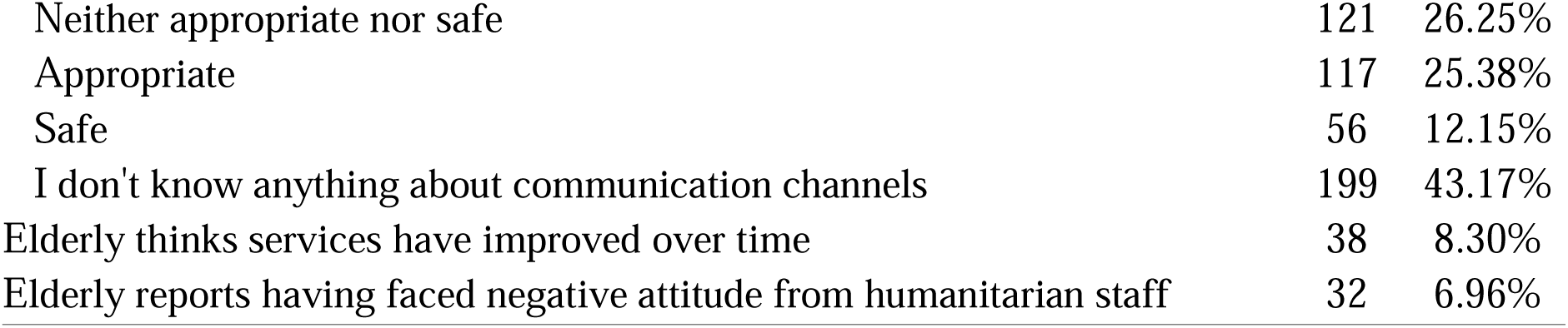
Receipt and assessment of humanitarian assistance.

|  | n | % |
| --- | --- | --- |
| Type of assistance received in the past two months |  |  |
| None | 64 | 13.88% |
| Multi-purpose cash assistance card | 128 | 27.77% |
| Cash-for-food assistance card | 323 | 70.07% |
| Winter cash for heating assistance | 241 | 52.28% |
| Food parcels/ assistance | 42 | 9.11% |
| Non-food assistance from charity | 10 | 2.17% |
| Covering healthcare expenses | 5 | 1.08% |
| Don't know | 6 | 1.30% |
| Has any humanitarian agency collected data on your needs since the COVID-19 crisis started in 2020? |  |  |
| No | 298 | 64.64% |
| Health | 137 | 29.72% |
| WASH | 36 | 7.81% |
| Protection | 5 | 1.08% |
| Food security and nutrition | 30 | 6.51% |
| Shelter | 16 | 3.47% |
| Other | 1 | 0.22% |
| How do you receive information about the humanitarian services? |  |  |
| SMS | 264 | 57.27% |
| Hotline | 5 | 1.08% |
| WhatsApp | 11 | 2.39% |
| Supermarkets | 4 | 0.87% |
| Posters and leaflets | 1 | 0.22% |
| Reception centers | 2 | 0.43% |
| Information centers in community centers | 5 | 1.08% |
| Distribution points | 4 | 0.87% |
| Municipality | 0 | 0.00% |
| Healthcare centers | 0 | 0.00% |
| Word of mouth (neighbors, relatives) | 29 | 6.29% |
| Visits from humanitarian agencies | 49 | 10.63% |
| Hasn't received any information | 91 | 19.74% |
| Other | 21 | 4.56% |
| Elderly faced problems in accessing information | 110 | 24.28% |
| Barriers towards accessing information about services |  |  |
| Financial situation | 10 | 8.06% |
| Connection to the internet | 9 | 7.26% |
| Being illiterate | 53 | 42.74% |
| Technology | 14 | 11.29% |
| Other | 54 | 43.55% |
| Facing difficulties in comprehending sent information? | 73 | 19.52% |
| Degree of satisfaction about humanitarian response |  |  |
| Very satisfied | 0 | 0.00% |
| Satisfied to a great extent | 13 | 2.82% |
| Satisfied to some extent | 214 | 46.42% |
| Not satisfied at all | 234 | 50.76% |
| Assessment of communication channels |  |  |
| Neither appropriate nor safe | 121 | 26.25% |
| Appropriate | 117 | 25.38% |
| Safe | 56 | 12.15% |
| I don't know anything about communication channels | 199 | 43.17% |
| Elderly thinks services have improved over time | 38 | 8.30% |
| Elderly reports having faced negative attitude from humanitarian staff | 32 | 6.96% |

#### Collecting Data on Needs

Most elderly (64.64%) reported never having had their needs assessed by any humanitarian agency. Less than a third of respondents (29.72%) reported ever being assessed for health needs, and smaller proportions were ever approached to assess needs in WASH (7.8%), protection (1.1%), food security and nutrition (6.5%), and shelter sectors (3.5%).

> No human has knocked on my door to ask how I’m doing. Organizations come and give money to other people, but nobody hears my voice.
>
> P10

#### Access to Information About Services

Around 20% of respondents reported not receiving any information about services from humanitarian agencies. This was significantly more common in hosted communities (31.2%) than in ITSs (18.3%) (p<0.001). The top perceived barrier was being illiterate (42.7%) and 43.2% reported not knowing anything about communication channels. Out of those who were aware of the communication channels, almost half (44.7%) considered them neither appropriate/easily accessible nor safe/posing no threat.

#### Overall Satisfaction

On a more global scale, when asked about the degree to which OSRs were satisfied with the humanitarian response, more than half reported not being satisfied at all (50.7%), and 46% reported being only satisfied to some extent. Only 13 participants reported being satisfied to a great extent. Higher dissatisfaction rates were reported by participants in hosted communities than ITSs (p=0.009).

> I feel there is no justice. They say: ‘Oh this lady, she lives in an apartment’ (not in a tent, so she must be doing well). Well, do you know how I’m living in this apartment? Do you know how I’m paying rent? I am a widow and I cannot leave the house to search for assistance and stand in lines.
>
> P13

Furthermore, 91.7% of the total sample considered there was no improvement in the services provided over time. This was also more significantly reported in hosted versus ITS residents (p<0.001). On the other hand, OSRs’ views on humanitarian field workers are positive with around 93% of OSRs reporting never having faced any negative attitude from humanitarian staff.

### Findings in the shelter sector

Table 4 shows the shelter and assets characteristics of the study sample.

**Table 4:** Summary of shelter and assets characteristics of the survey sample.

|  | n | % |
| --- | --- | --- |
| Number of people living in the same shelter (median; IQR) | 6 | 17 |
| Number of rooms in the shelter (median; IQR) | 2 | 1 |
| Number of hours of electricity | 9 | 15 |
| Currently under eviction notice | 65 | 14.1% |
| Heating sources |  |  |
| Electric heater | 4 | 0.9% |
| Diesel heater | 251 | 54.4% |
| Charcoal | 4 | 0.9% |
| Blankets | 301 | 65.3% |
| Gaz (methane) heater | 9 | 2.0% |
| Assets |  |  |
| Mattress | 284 | 61.6% |
| Blanket | 243 | 52.7% |
| Full set of winter clothing | 98 | 21.3% |
| Bed | 37 | 8.0% |
| Mobile phone | 253 | 54.9% |
| Phone connected to the internet | 225 | 88.9% |
| Problems in shelter |  |  |
| Inappropriate for living | 214 | 46.4% |
| Unsafe | 9 | 2.0% |
| Other | 11 | 2.4% |
| No problems | 236 | 51.2% |
| Contacted humanitarian agencies for assistance about shelter problem | 88 | 39.1% |
| Humanitarian agencies responsive to shelter issue | 8 | 9.1% |
| Barriers to contacting humanitarian agencies to fix shelter |  |  |
| Don't know whom and how to contact | 112 | 80.58% |
| Irresponsive staff | 25 | 17.99% |
| Other | 2 | 1.44% |

#### Issues and Unmet Needs

When asked about the quality of their shelter, 51.5% of ITS residents reported the shelter to be inappropriate for living, compared to 32.8% of hosted communities. The main reasons that rendered their shelter inappropriate were being ‘very cold’ (P02, P13), ‘leaking rainwater’ (P02, P05), ‘has mold formations on the wall’ (P07), posing health risks to the elderly residents. Only nine OSRs in the quantitative survey considered their shelter unsafe from a security perspective.

Half of the respondents reported less than 9 hours of electricity per day. The most common heating source was blankets (65.3%), followed by diesel heater (54.4%). Around 42.3% reported that blankets were their only source for heating.

Only 14% of elderly reported living under an eviction notice by the landlord, with the rate ranging between 9.6% in residential setting to 18.8% in ITSs. Per governorate, the highest reported eviction rate was reported in Bekaa (20.4%), followed by North and Akkar at 5.2% and 3.8% respectively.

Qualitative consultations have highlighted the issue of high rents. Even for those residing in tents, the rent cost is burdensome and may vary substantially from one location to another. For instance, P07 reported 750,000 LBP (34 USD) rent and electricity fees per month, compared to 500,000 (22.7 USD) paid by P09 living in two camps only a few kilometers apart. Hosted participants pay significantly higher rents; P12, P13, and P14 residing in a residential compound in Bekaa pay 1 million LBP (45.45 USD).

#### Humanitarian Assistance and Inclusion

Out of those who reported problems in shelter conditions, only around 40% contacted humanitarian agencies for assistance. Among those, only 8 participants (9%) reported humanitarian agencies to be responsive to the complaint. Out of the majority that did not contact agencies, the most commonly reported barrier was not knowing whom and how to contact (82%), and around 18% complained about the irresponsiveness of the staff. In addition, 52.3% of survey respondents reported receiving winter cash assistance for heating. Interviewed participants revealed that one form of humanitarian assistance was the provision of heaters and diesel, as well as blankets as part of the winterization assistance for residents of ITSs, particularly those located in areas highly prone to low temperatures and storms.

> They gave me those two blankets and two mattresses. I did not have those mattresses before. […] I put the mattress on the floor and sleep on it.
>
> P01

Interviewed participants reported that there was humanitarian provision of canvas to strengthen and support tents.

> The tent used to leak rainwater at the beginning of the inter. They gave us canvas, [the camp chief] gave it to me and the kids installed it. We used to be visible to the street, they installed it so I don’t show, because as you see, my tent is right on the asphalt (street).
>
> P05

In addition, participants reported receiving diesel heaters and diesel as part of the winter assistance (P02, P04 P05).

#### Coping Strategies

The high cost of rent forces OSRs to prioritize it over other necessities. For P07, the only source of income is the food cash assistance provided by the World Food Program (WFP), amounting to “1,800,000 LBP (81.81 USD)” transferable amount to a card per month for his entire family of six. The card is used at check-out at contracted supermarkets. Given the high rent and electricity cost, it is impossible to secure adequate food:

> The local grocery store just informed me I cannot buy on-credit any more. Now if I have to pay the rent of the caravan, and I have my son, he cannot pay the rent of his so I pay it for him, that’s 600[000], and say there’s 500[000] electricity bill. What remains in my card? They are right to stop giving me food on credit, they also want to work. […] Look at our fridge. There are only a few dates, and we didn’t even buy them they were distributed by a charity.
>
> P07

In addition, P13 reported that she “had gold rings and bracelets and had to sell them to pay rents since [they] migrated”. As for coping with the cold weather in light of the high diesel prices, she reported trimming old clothes and using them as firewood to warm up.

> We have these blankets, the UN handed them over to us a long time ago. I told my daughter-in-law to sort whatever we have to give away in the summer; let’s burn them now [to get warm]. Jackets, clothes, I swear, two blankets, I trimmed them into small pieced and we have been using them as firewood for the past four days.
>
> P13

Others resorted to selling diesel heaters and buying methane heaters to reduce consumption of high diesel (P02). Furthermore, it was common for OSRs to ration usage of the diesel heater and rely solely on blankets:

> We have not bought a liter of diesel this winter. They gave us diesel maybe 3 times here. Once with 20 liters and twice with 10 liters each. We live with that. For example, we do not turn it on until it’s dark. In the day, we cover up with blankets, because we cannot afford more diesel. […] in the evening we turn it on a little, we turn up the heat for a bit then we turn it off for 10-15 minutes until it’s cold again and we turn it on. This is how we do, what else could we do?
>
> P04

Finally, the provision of canvas without installation, especially for OSRs living alone, forces them to install the canvas themselves, posing high risk of injury.

> It was raining and the caravan was leaking. I climbed up to put something to stop the leakage, and it was windy, […] I fell from the caravan to the ground. On the cement.
>
> […]. Afterwards I found myself failing to get up. I deteriorated until I found myself on the wheelchair.
>
> P08

### Findings in the health sector

Table 5 displays the health characteristics of the survey participants.

**Table 5:** Summary health characteristics of the survey sample.

|  | n | % |
| --- | --- | --- |
| Bodily pain over past 4 weeks |  |  |
| None | 48 | 10.41% |
| Very mild | 15 | 3.25% |
| Mild | 65 | 14.10% |
| Moderate | 160 | 34.71% |
| Severe | 173 | 37.53% |
| Disability |  |  |
| Has difficulty seeing, even with eyeglasses | 310 | 67.25% |
| Has difficulty hearing, even with hearing device | 135 | 29.28% |
| Has difficulty walking or climbing stairs | 313 | 67.90% |
| Has difficulty bathing | 120 | 26.03% |
| Has difficulty putting on and taking off clothes | 101 | 21.91% |
| Has difficulty using the bathroom | 96 | 20.82% |
| Has difficulty sitting or getting off a chair or bed | 206 | 44.69% |
| Has difficulty continence | 94 | 20.39% |
| Has difficulty eating | 91 | 19.74% |
| Has difficulty taking care of appearance (combing, grooming) | 71 | 15.40% |
| Has difficulty using the phone | 99 | 33.33% |
| Has difficulty using transportation | 115 | 28.54% |
| Has difficulty shopping | 93 | 25.62% |
| Has difficulty preparing meals | 92 | 37.55% |
| Has difficulty doing housework (laundry, cleaning, ...) | 138 | 59.48% |
| Has difficulty managing and taking medications on time | 83 | 19.86% |
| Has difficulty remembering and concentrating | 161 | 34.92% |
| Needs a caregiver | 292 | 63.34% |
| Caregiver unavailable | 42 | 14.38% |
| Need for assistive devices |  |  |
| Dentures | 224 | 48.59% |
| Cane/ walking stick | 89 | 19.31% |
| Crutches | 29 | 6.29% |
| Eyeglasses | 281 | 60.95% |
| Hearing device | 87 | 18.87% |
| Wheelchair | 17 | 3.69% |
| Diapers | 14 | 3.04% |
| Chronic Diseases |  |  |
| Diagnosed chronic diseases |  |  |
| None | 56 | 12.15% |
| Osteoporosis | 31 | 6.72% |
| Disc issues | 112 | 24.30% |
| Hypertension | 197 | 42.73% |
| Diabetes | 105 | 22.78% |
| Hypercholesterolemia | 57 | 12.36% |
| Cardiovascular Disease | 109 | 23.64% |
| Chronic respiratory diseases (COPD, Asthma) | 59 | 12.80% |
| Rheumatoid Arthritis | 48 | 10.41% |
| Kidney disease | 25 | 5.42% |
| Cancer | 4 | 0.87% |
| GI diseases (ulcer) | 76 | 16.49% |
| Neurological diseases | 40 | 8.68% |
| Other | 93 | 20.17% |
| Don't know/ refuse to answer | 3 | 0.65% |
| Number of chronic diseases |  |  |
| None | 55 | 12.01% |
| One | 120 | 26.2% |
| Two | 122 | 26.64% |
| More than two | 161 | 35.15% |
| Medication Practices |  |  |
| Elderly prescribed medications | 349 | 86.60% |
| Adherence to medication regimen |  |  |
| I don't take my meds at all | 38 | 10.89% |
| I take my meds as prescribed | 246 | 70.49% |
| I take my meds but not as prescribed | 65 | 18.62% |
| Means to get medication |  |  |
| Out-of-pocket money | 219 | 70.42% |
| Subsidized/ at no cost at dispensaries | 68 | 21.86% |
| Donations from charity | 18 | 5.79% |
| Other | 6 | 1.93% |
| Elderly resorts to reducing or selling food intake to buy meds | 174 | 56.49% |
| Reasons for non-adherence to medication regimen |  |  |
| Medication unavailable | 49 | 47.57% |
| Medication expensive | 94 | 91.26% |
| Medication triggered side effects | 3 | 2.91% |
| Felt better so stopped taking the medication | 1 | 0.97% |
| Distance and transportation to reach pharmacy/ dispensary | 2 | 1.94% |
| Physical barrier/ unable to go get meds and no one to help | 4 | 3.88% |
| Used to get meds from Syria | 7 | 6.80% |
| Access to Primary Healthcare |  |  |
| How many times did you need primary healthcare during the past 6 months? (Median, IQR) | 2 | 4 |
| Were you able to access this needed healthcare the last time you needed it? | 244 | 78.96% |
| Barriers towards accessing primary healthcare the last time it was needed |  |  |
| Distance of health center | 9 | 13.85% |
| Transportation cost | 30 | 46.15% |
| Physical disability limiting access the health center | 1 | 1.54% |
| Inadequate welcoming/treatment by health center staff | 5 | 7.69% |
| Fees doctor visit | 28 | 43.08% |
| Cost of drugs/Diagnostic tests/treatment | 37 | 56.92% |
| Not accepted | 2 | 3.08% |
| Doesn't know where to go | 1 | 1.54% |
| Long waiting time | 8 | 12.31% |
| Other | 3 | 4.62% |
| Did you pay for the primary health care assistance? |  |  |
| No, totally free | 47 | 19.18% |
| Yes, but discounted/subsidized/financial contribution/ cost sharing | 131 | 53.47% |
| Yes, household needed to pay in full | 66 | 26.94% |
| Health Inclusion Indicators |  |  |
| Elderly reports available of healthcare services for all their conditions and needs | 289 | 62.69% |
| Elderly reports affordable healthcare services (including medications) | 167 | 36.54% |
| Elderly thinks provided healthcare services are of good quality | 329 | 76.33% |
| Elderly thinks they are provided healthcare services on equal basis to others | 314 | 69.93% |
| Elderly reports presence of health services by humanitarian agencies that reach them at home if they cannot make it to the healthcare facility | 23 | 5.78% |
| Elderly reports that healthcare providers take their informed consent before intervening | 398 | 88.84% |
| Elderly reports interrupted care | 148 | 32.10% |

#### Issues And Unmet Needs

Pain was found to be a significant issue in OSRs; around 37% of participants reported severe pain, and 34.7% reported moderate pain, while only 10.4% reported not feeling any pain over the past 4 weeks. Females suffered higher pain levels (p<0.001). Indeed, bodily pain was reported by five out of eight female OSRs interviewed in the qualitative consultations; it either manifested as a symptom of diagnosed chronic conditions or as a result of disability.

> Everywhere hurts. My arms all the way to my hands, my feet, they all hurt. My shoulders hurt as if they’re perforated with a bradawl.
>
> P03

At the level of functional status. most survey participants (63%) indicated that their health condition impeded their capacity to engage in work or carry out daily household responsibilities. Among them, a substantial proportion (87.2%) reported this hindrance persisted for a duration exceeding three months. The overwhelming majority of respondents (93.3%) reported having difficulty performing at least one ADL or IADL, and 74.6% reported difficulty performing two or more. Specifically, 67.2% of respondents reported difficulty seeing even with eyeglasses, 29.3% reported difficulty hearing even with a hearing device, 67.9% reported difficulty walking or climbing stairs, 26% reported difficulty bathing, and 21.9% reported difficulty putting on and taking off clothes. Moreover, 20.8% reported difficulty using the bathroom, 44.7% reported difficulty sitting or getting off a chair or bed, 19.7% reported difficulty eating, 15.4% reported difficulty taking care of appearance such as combing and grooming, and 33.3% reported difficulty using the phone. In addition, 28.5% reported difficulty using transportation, 25.6% reported difficulty shopping, 37.6% reported difficulty preparing meals, and 59.5% reported difficulty doing housework such as laundry and cleaning. Lastly, 19.9% reported difficulty managing and taking medications on time, and 34.9% reported difficulty remembering and concentrating (Table 5).

Among those interviewed in the qualitative consultations, 7 were living with at least one type of functional difficulty significantly affecting their quality of life. One participant also had to take care of her paraplegic spouse:

> He shouts at me and I’m ill. […] He wakes up very early, and I need to wake up with him. Whether I’m good or ill. I came back from my surgery and changed his diaper.
>
> P11

With respect to the need for assistive devices, 292 respondents (63.34%) reported needing a caregiver out of whom 14.38% had no caregiver available. Females were significantly more likely to need care compared to males, even after controlling for age (p<0.001). Nearly half OSRs (48.59%), two-thirds (60.95%) and 18.87% reported needing dentures, eyeglasses, and hearing aids, respectively. None of the participants in the qualitative consultations were aware of an agency providing assistive devices for elderly.

The most significant health burden was found to be chronic diseases. A total of 403 (87.4%) OSRs reported being diagnosed with at least one chronic illness, and a comorbidity rate of 61.4%. Hypertension (HT) was the most frequently reported disease (42.73%) followed by disc problems (24.30%), diabetes (22.78%), and cardiovascular disease (23.64%). Other conditions that were reported by a smaller proportion of respondents included respiratory diseases such as COPD and asthma, rheumatoid arthritis, kidney disease, gastrointestinal diseases such as ulcer, neurological diseases, and cancer. Females disproportionately suffer more from diabetes (p=0.001), hypertension (p<0.0001), and have more comorbidities than males (p<0.0001), even after controlling for age. Among the 14 participants in the qualitative component, 10 reported having at least one chronic disease. Conditions include diabetes (P01, P08, P13, P10), HT (P01, P02, P06, P13, P14), chronic pain (P03, P02, P05), heart disease (P04, P06), disc problems (P02), rheumatism (P05), and stomach ulcer (P11).

#### Humanitarian Services and Inclusion

First, humanitarian health actors provide chronic medications for no or subsidized cost at contracted Primary Healthcare Centers (PHCCs) and dispensaries. Out of those diagnosed with at least one chronic illness, 86.6% reported being prescribed medications for their condition. Among those, 70.5% adhered to their regimens, 18.6% took them inconsistently, and 10.9% (38) didn’t take them at all. The most common reason for nonadherence was medication cost, reported by 91.3% of respondents. Other reasons for nonadherence included medication unavailability (47.6%), and physical barriers preventing access to medication (3.9%). Very few respondents reported stopping medication because they felt better or experienced side effects. The most common means of obtaining medications was out-of-pocket money, reported by 70.4% of respondents. 21.9% reported obtaining medication through subsidized or no-cost dispensaries, while smaller proportions reported obtaining medication through donations from charity (5.8%). Qualitative findings affirmed these insights and shed light on available medication services for refugees and the emerging challenges. While some funded dispensaries offer chronic medications for a minimal cost or even free of charge, the economic crisis has severely affected medication availability.

> PHCC [X] have two medications on one day, three on another, one on another. With Doctors Without Borders, we were more content (in reference to the support this INGO had provided to the PHCC)
>
> P04

Some types of chronic medications were either not available in dispensaries, or not available in the country altogether. For those available outside dispensaries, the cost burden prevented OSRs from purchasing them or adhering to their treatment regimens. They reported “[buying] only one blister pack because [they] could not afford the entire pack of medication” (P02), and “[she] no longer has money so [she] stopped buying the painkiller and any other medication” (P03). Transportation is also a burden to getting medications. P05 reported that she “sometimes send[s her teen son] to go get them on foot, so save the 20[000 LBP cost of transportation]”.

Second, at PHCCs supported by humanitarian (I)NGOs, medical consultations are subsidized, costing 3,000 LBP to 5,000 LBP (0.14 USD to 0.23 USD, at the time of data collection). However, a significant concern was voiced when refugees sought medical assistance at non-supported PHCCs because these were geographically inaccessible, or because they mistrusted the services at nearby supported PHCCs due to perceived bad quality of care. This sometimes forced them to seek care in private clinics, imposing a heavy financial burden.

> We need to see a doctor. We need to pay, and we don’t have enough money. I swear he takes 450,000 LBP (21 USD). I don’t go to the PHCC, my daughter, [because] they don’t know how to treat me. I have diabetes, hypertension, and stomach pain. I am scared of taking the wrong medication.
>
> P01

A total of 309 (67%) participants reported needing PHC at least once over the past 6 months. Of those, 244 (79%) reported being able to access the needed care the last time they needed it. Barriers to accessing PHC among those were primarily attributed to the cost of drugs/diagnostic tests/treatment (56.9%), transportation cost (46.2%), fees for doctor visits (43.1%), and the distance of the health center (13.8%). Most respondents (53.47%) reported paying for primary health care services, albeit at discounted rates. Only 19.18% received the care free of charge. On the other hand, 26.94% of participants indicated that they had to make full payments for the services.

> They take 3,000 and sometimes 5,000 LBP […]. My cardiologist and dentist there are very good, and the ladies (nurses) at the PHCC are very respectful.
>
> P05

##### • Access to Hospitalization

While UNHCR supports up to 75% of the cost for life-saving emergencies and other elective medical interventions by approval of its Exceptional Care Committee, the remaining financial burden remained substantial. Additionally, for geographically accessible hospitals not contracted by UNHCR, refugees were forced to pay completely out-of-pocket, posing a tremendous financial burden. Moreover, bureaucratic delays from the committee in approving surgery cost coverage, might be associated with irreversible and devastating consequences on OSRs’ health. P08 shared his experience needing spine surgery after a domestic accident trying to fix his tent:

> I told the doctor we cannot afford [the surgery]. He said you can apply to UNHCR to cover the expenses. I applied. They got back to us after almost one and a half months. At this point my situation deteriorated a lot. After two months they informed us that the request is rejected and asked for a neurography. The doctor told me I must do the surgery. He applied again for it; it also took them a month to reply. By that time, I was done, I needed someone to lift me.
>
> P08

Considering access to hospitalization, only 87 participants (18.9%) reported needing hospitalization in the past 6 months, out of whom 23 (73.6%) were able to access it the last time they needed it. Seven respondents (10.94%) received hospital care totally free of charge, while 34.3% indicated receiving financial contribution from UNHCR. However, almost 40% reported having had to pay totally out-of-pocket for hospital care. The most prevalent barriers to hospitalization were financial (61%), followed by transportation cost (39%).

Finally, while laboratory and diagnostic tests were up to 85% covered by UNHCR at supported PHCCs at the time of the study, qualitative interviews showed that medical images posed a significant financial burden. The lack of financial assistance in this area added to the overall struggle faced by elderly refugees in obtaining necessary medical diagnostic services, especially for those who were unable to afford the remaining 15%.

> [The lab results of my stomach ulcer biopsy] are out. I could not go get them; I don’t have the money for them. They cost around 80,000-90,0000 LBP (3.6 to 4 USD)
>
> P11

#### Indicators of Inclusion in Health Services

A total of seven indicators were measured to describe several dimensions of health services inclusivity of OSRs. First, relevance and comprehensiveness of services: Participants were asked whether there are healthcare services available for all of their conditions and needs. Most of participants (62.69%) responded affirmatively, while 22.78% denied, and 14.53% did not know. Second, financial accessibility or affordability: The majority of participants (63.46%) responded that the services were not affordable. Third, reflects OSRs’ assessment of the quality of the healthcare services provided. The majority of participants (76.33%) reported that the services were of good quality. Fourth, perception of equality in receiving services. Almost 70% responded that they were provided with healthcare services on an equal basis compared to others. Fifth, this indicator tackles the dimension of physical inclusivity for home-bound elderly. A significant majority of participants (81.34%) responded that there were no healthcare services provided by humanitarian agencies that reach the respondents at home if they cannot make it to a healthcare facility, while 13.67% responded that they did not know it existed. Sixth indicator examines the ethical dimension of provided health services in terms of respecting the elderly’s autonomy through informed consent. The majority of participants (88.84%) responded that healthcare providers explain what they will do and take the respondents’ permission before their intervention. Seventh and last indicator assesses the continuity of care. Half the respondents (50.5%) reported having their care continued, while a third (32.1%) reported their care was interrupted when they should have been referred to other specialists or testing, and 17% reported not knowing the answer.

#### Coping Strategies

OSRs resort to a number of strategies to cope with the unavailability and high cost of medications. First, 56.4% reported resorting to reducing food intake or selling food assistance to buy medications. They may also use their food cash assistance to buy the medications, reducing the budget allocated for other needs:

> I pay all the money in my card to buy medication. If it wasn’t for the cost of medication I would have buy [more] food.
>
> P01
>
> I get three medications out-of-pocket. Every month I pay 500,000 LBP; the amount of the card (food cash assistance) that I receive.
>
> P05

Furthermore, OSRs incur debt for medication (P01), hospitalization (P11), and medical imaging (P08). Finally, 6.8% of survey participants reported obtaining medications from Syria.

> I do not take my [prescribed] medications. When they get them for me from Syria, I take them.
>
> P10

### Findings in nutrition and food security

Table 6 summarizes the food consumption and coping strategies of the survey sample.

**Table 6:**
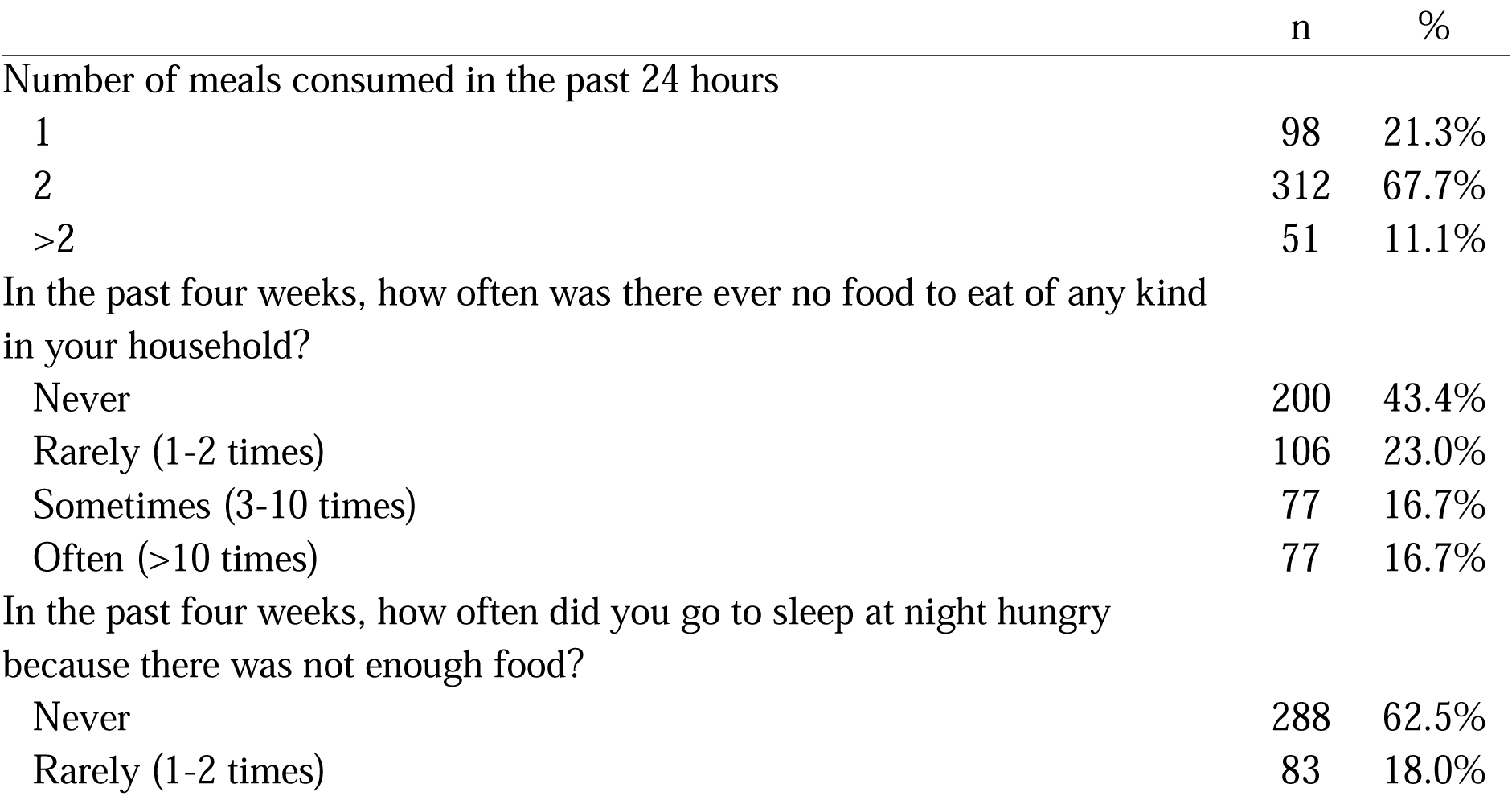
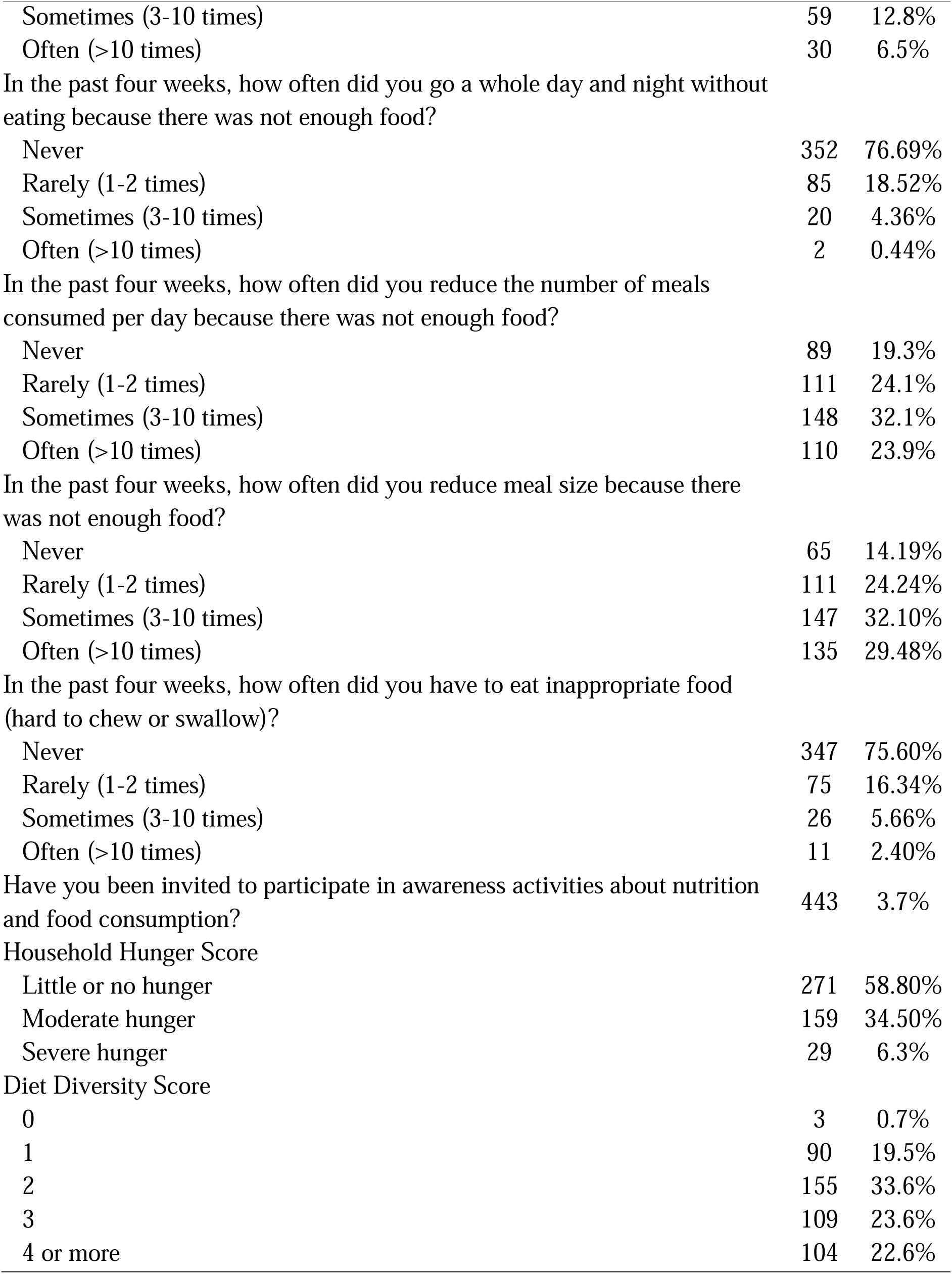
Summary of food consumption and food coping strategies of the survey sample.

#### Issues and Unmet Needs

The majority of respondents (79%) have consumed 2 meals per day in the past 24 hours to the survey. We also used the diet diversity to inspect the consumption of a total of 11 food groups: milk and milk products, cereals, roots and tubers, fruits and vegetables, meat products, eggs, fish, legumes, nuts and seeds, fats, sugar, and other. A total of 21% have consumed 0 or 1 food group, a third consumed 2 food groups, and 22.5% consumed more than 2 groups. In terms of nutrient groups, OSRs mostly consumed Vitamin B complex (64.6%), all-source protein (45.12%), and all source vitamin A-rich foods (44%). The least consumed groups include iron-rich foods (15.84%), zinc-rich foods (20.8%), and calcium-rich foods (23.4%).

#### Humanitarian Services and Inclusion

The most important food assistance provided is the large-scale distribution of cash-for-food assistance via an electronic card monthly transfer by the WFP. This type of assistance allows Syrian refugees to either withdraw their money from ATMs or use the card directly at contracted supermarkets. The amount of assistance provided depends on the number of household members at the time this study was conducted. This assistance is received by 70% of the survey participants, and is deemed crucial but inadequate for accessing food and other basic needs.

> [The food assistance] was 40[000] LBP (equivalent to 26.7 USD per person per month). Now it’s 300[000] LBP (equivalent to 13.6 USD per person per month). It is 300[000], yes but it doesn’t do anything at all with the high prices. It has been 3 months now that my son asks me to get one kilogram of chicken thighs to make *shawarma*.
>
> P05

The distribution of food parcels has been infrequent since the beginning of the economic crisis and the COVID-19 pandemic; only 9.1% reported receiving food assistance in the past two months.

> The last time [we received food parcels] was last Ramadan. We received 3 parcels in Ramadan then none. So, it has been almost a year.
>
> P08

and only a tiny percentage (3.7%) of respondents reported being invited to participate in awareness activities about nutrition, none of which are specifically targeting older people. None of the participants in the qualitative interviews reported accessing any nutrition awareness services.

#### Coping Strategies

Results of the household hunger scale indicate that the majority (59%) of respondents experienced little or no hunger, while a third (34.6%) experienced moderate hunger, and only 6.3% experienced severe hunger. The most commonly experienced negative food coping strategies include having to reduce the number of meals consumed per day, and reducing meal size, experienced “sometimes” or “often” in 56% and 61.2% of the cases, respectively. Qualitative consultations echoed this finding and showed the frequency and severity of reducing food quantities:

> As for eating and drinking, for instance if I want to cook today, I calculate how much I can afford food per day given the monthly budget. Let’s say 40,000-50,000 LBP (∼ 2-3 USD). Then I must eat as little as that, because if I eat more, I will have to borrow money, and if I do, I won’t be able to pay it back.
>
> P04
>
> How do we spend? As the saying goes: eat so you don’t die. […] Instead of eating two pieces of bread, we eat half a piece or three quarters of a piece.
>
> P06

Other participants mentioned resorting to selling some items in the food parcel they receive to buy their food needs (P04).

### Findings in water, sanitation, and hygiene

Table 7 summarizes the WASH assessment responses by the survey participants.

**Table 7:**
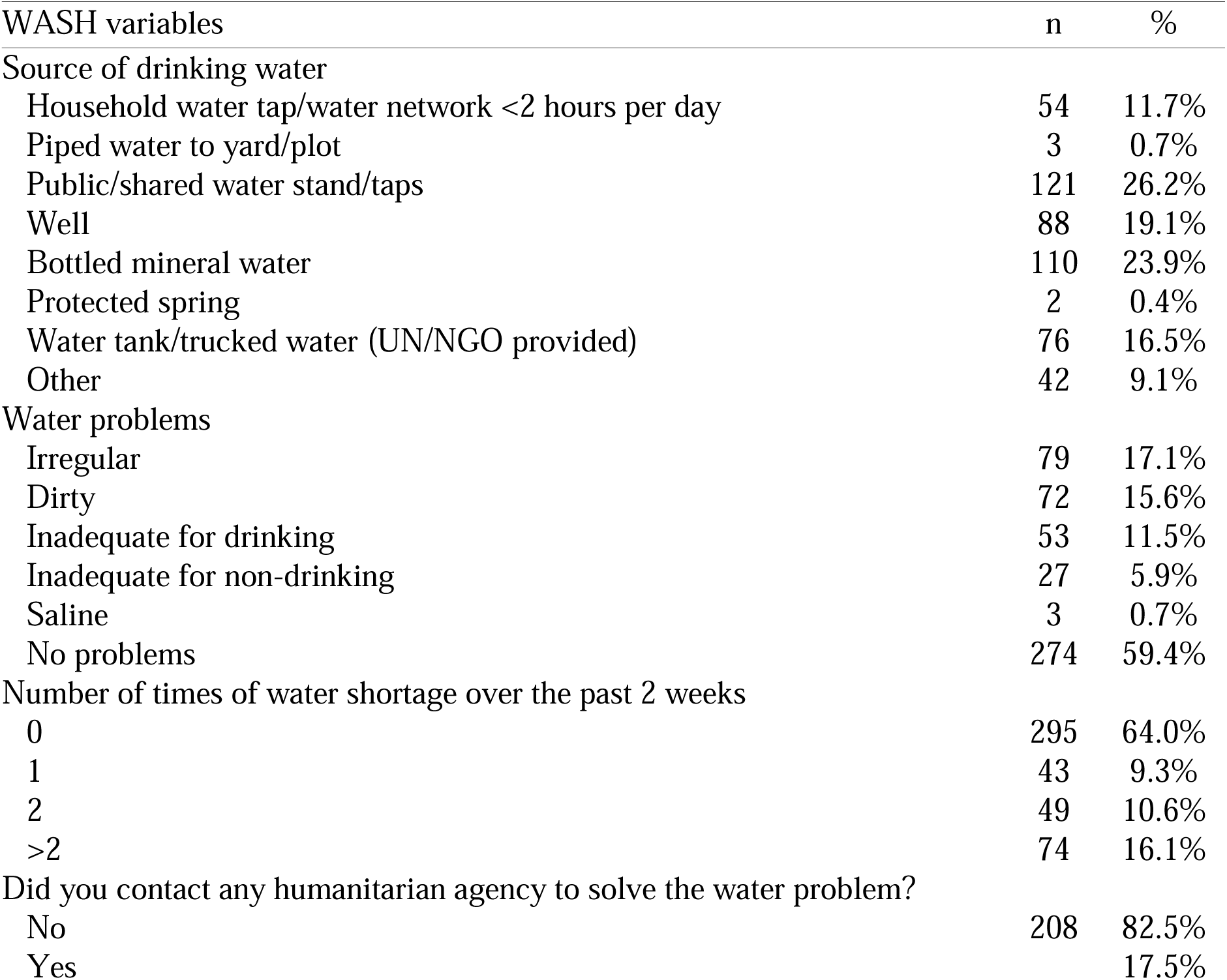
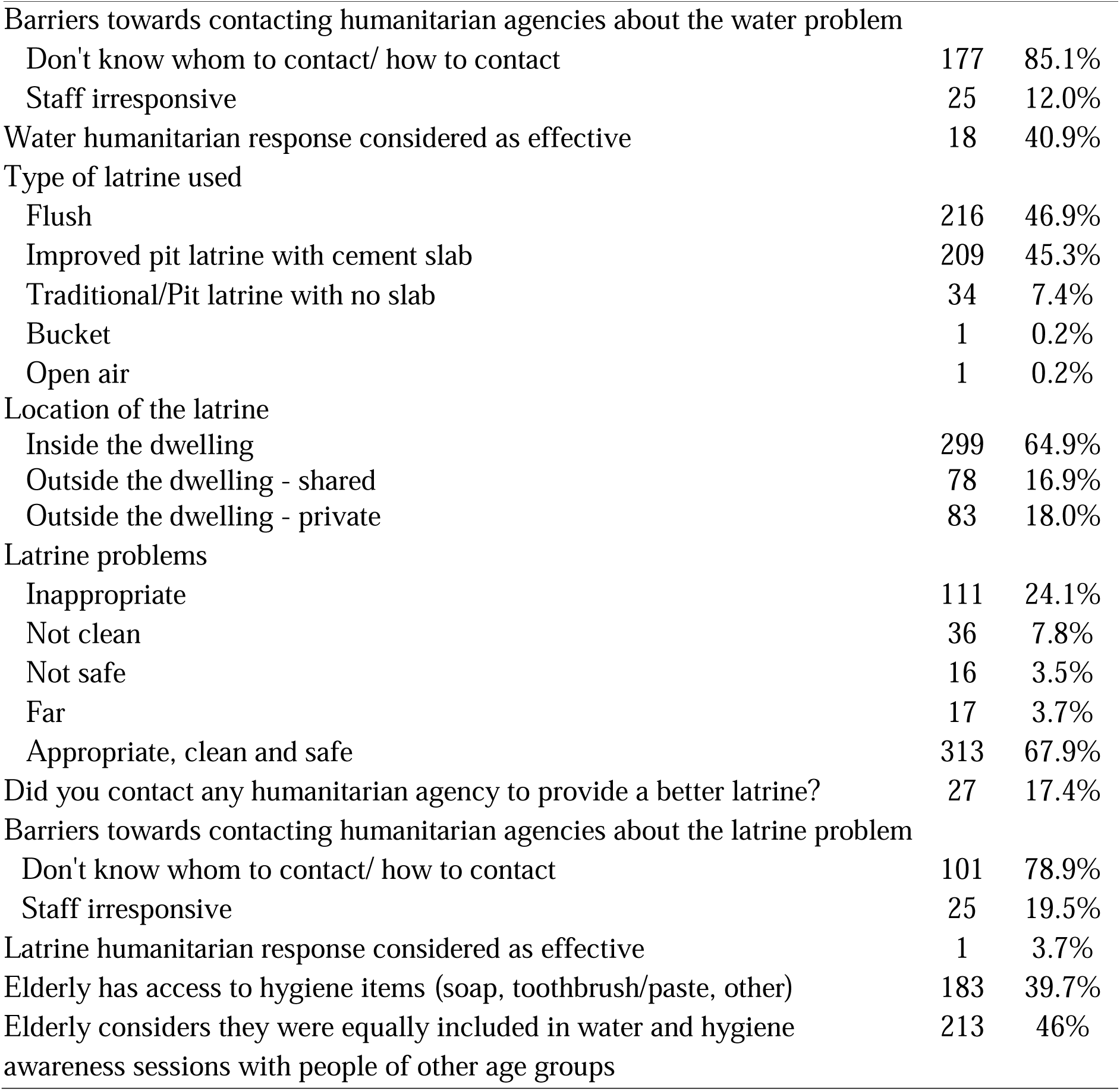
Summary of water, sanitation, and hygiene assessment of the survey sample.

#### Humanitarian Assistance and Unmet Needs

In ITSs, humanitarian agencies acting in the WASH sector install and maintain water and sanitation facilities in coordination with the Ministry of Power and Water. Activities include water trucking and wastewater desludging. On the other hand, refugees in hosted communities benefit from WASH services similar to Lebanese hosting populations, which are mainly provided by the water establishments. Humanitarian NGOs also support water establishments maintain consistent and reliable access to water through minor repairs and operational support, as well as rehabilitation and maintenance of wastewater treatment plants and related infrastructure [17].

In regards to access to water, q quarter of respondents (26.2%) relied on public/shared water taps, followed by bottled mineral water (23.9%), well water (19.1%), and water tank/trucked water (16.5%). Notably, 59.4% reported having no water problems, while irregularity (17.1%) and dirtiness (15.6%) were the most common issues. Concerning water shortages, the majority (64%) experienced no shortage in the past two weeks, while 16.1% faced shortages more than twice. Only 17.5% of respondents who reported water problems reached out to humanitarian agencies for assistance because of uncertainty about whom to contact/how to contact (85.1%) and unresponsive staff (12%).

> No, thank God. Water does not cut; there’s a lot of water.
>
> P01

In terms of latrines, flush toilets (46.9%) and improved pit latrines with a concrete slab (45.3%) were prevalent. The majority (64.9%) had their latrines located inside their dwellings, while 18% used shared external latrines. Notably, most OSRs considered latrines appropriate, clean, and safe (67.9%). Only a small proportion (17.4%) of respondents contacted humanitarian agencies for latrine improvement. The main obstacles reported were uncertainty about whom to contact/how to contact (78.9%) and unresponsive staff (19.5%). Shared toilets in some of the bigger ITSs were a controversial issue. While some participants had access to relatively well-maintained facilities, others highlighted poor hygiene due to over-crowdedness:

> 10 to 12 families share a bathroom. What hygiene would you find? If one woman takes her son to the bathroom you will have to wash the toilet to [use] it afterwards. We used to live in a house with two bathrooms. Now we live in a camp so we must bear.
>
> P05

In addition, shared toilets in relatively big tented settlements pose accessibility challenges for elderly with disability, because these facilities are usually located at the periphery of the settlement.

> If I need to go to the bathroom, I feel embarrassed of the people I force myself to walk (and not crawl). I stop and rest on my way to the bathroom. My back hurts a lot – I have two discs, I have rheumatism, my feet hurt.
>
> P02

As for access to bathing facilities, many elderly refugees, especially in ITSs, faced challenges when it came to showering in shared facilities during the winter due to the cold weather, as opposed to a warmer, more private ambiance when the bathing facility was inside the tent.

> I take my baths there, yes, when the weather allows such as today. I bathed yesterday. I mean […] in winter we bathe less. It’s too cold and windy, we can’t.
>
> P02

With regards to access to hygiene items, approximately 40% of respondents had access to hygiene items such as soap, dish washing liquid, and cleaning detergents. Only one participant in the qualitative consultations mentioned this type of assistance, mentioning they were given “these masks for Corona (COVID-19)” (P01). In terms of inclusion in participation in water and hygiene awareness sessions, more than half (52%) did not feel equally included with other age groups.

#### Coping strategies

More than a quarter of OSRs reported fetching water themselves (28.4%). This practice imposes physical, financial, and safety burdens on them.

> They pump water out of the well [for us in the camp]. No one can drink it or use it except for the bathroom. We have just filled those 8 plastic gallons for 20,000 LBP (∼1 USD) from the water tank. (…) I don’t drink from this water; I go fill from a charity water tap. I take the empty gallons and fill them, then come back on a passerby’s motorcycle.
>
> P09

Cold and hardly accessible shared bathing facilities outside tents pushes some elderly to bathe less frequently, and inside the tent, especially during the winter:

> During the summer we bathe in the bathrooms, but not in the winter. I am scared to go bathe there. I would fall very ill if I get cold. So, I heat some water [inside the tent] and bathe here. The water flows like that (outside the tent).
>
> P01

## Discussion

The current study presents the first extensive insights into unmet needs and humanitarian inclusion indicators of elderly refugees as reported by OSRs in Lebanon across the shelter, health, nutrition and food security, and WASH sectors. It used a mixed-methods design which enriched the findings from each strand by the other. Quantitative findings mainly shed light on unmet needs and pre-defined indicators of humanitarian inclusion, while qualitative results revealed perceptions, emotions, and suffering associated with these needs, as well as coping mechanisms not solicited in the quantitative survey, and issues with the humanitarian response. Integrated results show that OSRs have significant sector-specific and cross-cutting unmet needs nested under a lack of inclusive humanitarian interventions.

### Issues, unmet needs, humanitarian assistance and coping strategies

At the level of shelter, slightly less than half of OSRs reported living in inappropriate homes, and the main complaints raised were high rents, rain leaks, and cold, where a significant proportion reported blankets as the only source for heating. Humanitarian agencies provide blankets, diesel heaters and fuel, and canvas to support tents and caravans. Unfortunately, these types of services are inadequately addressing needs, pushing refugees to prioritize rent over other necessities such as food, and reducing heater usage.

At the level of health, the economic collapse in Lebanon has led to a rise in the cost of healthcare services and the lack of a universal and complete healthcare coverage, making them increasingly unaffordable for OSRs. The cost of transportation to reach healthcare facilities was another considerable barrier, particularly with the soaring prices of fuel and gas. These financial barriers limited access to essential medical services, exacerbating the health challenges faced by OSRs. Indeed, a striking majority of OSRs reported suffering from bodily pain. Furthermore, the majority reported having some type of disability or difficulty to perform daily tasks, and more than half indicated needing a caregiver. Not surprisingly, 87% of OSRs suffer from at least one NCD, mainly hypertension, disc problems, and diabetes. The direst need reported in this sector is for medications that were either unaffordable or unavailable. In addition, OSRs could not afford undergoing any surgical procedure when not covered by UNHCR or health sector partners, without going into debt. This posed risk on their lives and wellbeing due to delays and neglect. Humanitarian assistance does provide coverage for some chronic medications, medical consultations, and hospitalization. However, this coverage is not universal, pushing a significant proportion of OSRs to partially or totally skip their treatment regimens, to incur debts, and to spend most of their cash assistance on medication at the expense of food [26, 33, 42]. Access to healthcare in Lebanon is faced with cost barriers for both Lebanese and Syrians with costs of services, medications, and transportation hindering access to care [17, 33, 43–45]. Findings in this sector are consistent with other studies on older refugees in Lebanon [23, 25, 26, 46] and Jordan [27], which show the high burden of NCDs and difficulties in accessing healthcare. In Ukraine, older refugees were also found to suffer from a high burden of NCDs [47], and around 34% are in urgent need of chronic medication [48]. These findings are unfortunately consistent across different disaster settings in LMICs, where older populations affected by disasters have compelling health needs but lack access to healthcare due to cost barriers [49].

At the level of nutrition and food security, most OSRs are poorly nourished with a low diet diversity score and more than a third belong to moderate hunger on the household hunger scale. Although some humanitarian assistance includes distribution of food parcels and food cash assistance by WFP, these don’t seem to cover needs given the lack of other sources of income and deteriorating economic conditions. Main food coping strategies with poverty included reducing number of meals, meal sizes, and relying on more affordable food types, consistent with other findings on Syrian refugees in Lebanon [26, 33, 42]. Furthermore, the survey showed that OSRs are not included in nutrition awareness programs. Given the delicate physiological status at this life stage, it is imperative that OSRs are able to access food that is clean, diverse, nutritious, compatible with existing chronic illnesses, and easy to chew and digest. Elderly refugees can be at high risk of malnutrition due to multiple risk factors related to ageing and the risks of forced displacement [7, 50–53].

Finally, at the level of WASH, some OSRs lacked access to affordable safe drinking water and appropriate latrines, especially when facilities are shared. Main issues included poor hygiene and privacy, as well as lack of accommodations for older people with physical disabilities. While humanitarian agencies have worked in establishing WASH networks at ITSs, ensuring water trucking and desludging of wastewater, more inclusive programming and design is needed. Providing appropriate and accessible WASH facilities is a fundamental need to older refugees who suffer from mobility loss and difficulty in maintaining self-care and hygiene.

### Humanitarian Inclusion Standards

A key objective of this study is to assess humanitarian inclusion of older refugees using a set of indicators mapped on five pre-selected HIS’s. We do not claim that these indicators are comprehensive, but we believe they were properly adapted to the context to give preliminary evidence on the inclusion status as reported by the OSRs themselves.

#### Inclusion standard 1: Identification

This standard is said to be achieved if older refugees are identified to ensure they access relevant and appropriate services. This requires that age-disaggregated data are collected at all stages of the response, analyzed and used to inform proper programming and implementation. When OSRs in this study were asked if they were ever assessed for their needs, two thirds of them reported never having had their needs assessed by any humanitarian agency.

#### Inclusion standard 2: Safe and equitable access

The second standard uses data from the first to identify barriers and enablers to accessing assistance. Ultimately, the goal is to address and resolve barriers, and to enhance existing enablers, while including target groups at all stages in a participatory manner. The study shows that OSRs are facing multiple barriers: *environmental* barriers such as physical barriers with shared latrines far from the elderly’s residence, and transportation costs to reach healthcare centers and services. In addition, there are information barriers related to availability of assistance and services. Second, *institutional* barriers seem to also be present, because no needs assessments or age-sensitive interventions have been done specifically with OSRs despite most of the sample being in camps that are highly reachable by humanitarian agencies. This reflects either a lack of a policy, or a lack of implementation of existing policies.

#### Inclusion standard 4: Knowledge and participation

Study findings show a lack of any type of approach to OSRs to inform them of their rights or and entitlements, and participate in decisions affecting their lives, such as to consult with them about designing services for them.

#### Inclusion standard 5: Feedback and complaints

The fifth standard assures safe and accessible feedback and complaints channels. The study showed that at the level of basic assistance, shelter, WASH, OSRs failure to reach out to humanitarian organizations for complaints or requests is due to their lack of knowledge about the “who” and “how”, as well as agencies’ unresponsiveness.

#### Inclusion standard 7: Learning

While the learning standard is best assessed through an evaluation of organizational processes, one subjective high-level indicator is whether improvement in the quality of the intended outcome is perceived to have occurred. Our findings show that more than 90% of OSRs reported no improvement of provided services to them over time.

These findings from OSRs’ perspective complement results from Abi Chahine’s recent study assessing inclusion from humanitarian actors’ perspectives [14], showing the other side of the same coin. It was evident that service providers are indeed blinded to the needs of OSRs, and that ageist perceptions at the micro-level of institutions “excluded [elderly refugees] from livelihood programs, vocational training and cash assistance because their age identified them as “unproductive””. This exclusion could stem from ageist stereotypes and beliefs, as well as gaps in knowledge, policies, and standard operating procedures at the level of humanitarian organizations. Hence, study findings further support the body of evidence on older people’s exclusion in humanitarian response [11, 12, 15, 16].

### Limitations

The study findings must be read in light of its limitations. While the study employs two strands for triangulation and expansion increasing its credibility, not all findings were generated from both strands; some findings, especially those on coping strategies were revealed rather inductively in the qualitative strand. In addition, the study uses non-probability sampling approaches which induces selection bias due to the non-representative nature of the sample. While this might affect the generalizability of our findings, it’s important to note that the settings we have visited are not representative of the overall ITS and hosted status in Lebanon; the sample was more biased towards better rather than worse living circumstances and humanitarian attention, especially the ITS sample included in the qualitative component which came from mainly two large ITSs. In addition, the hosted community sample in both the survey and the qualitative component are smaller in size and may also be biased to more convenient and accessible sites. Hence, we claim that our study presents a relatively polished depiction of the reality of older refugees in Lebanon. Second, due to logistical reasons, we were not able to include qualitative insights from older refugees in hosted settings from the North governorate. However, we believe that insights shared by other participants are generalizable. Finally, the presence of measurement bias stemming from the interaction between interviewers and interviewees is possible. Variations in spoken dialects could have introduced inconsistencies in the data collection process. However, efforts were made to mitigate these biases through rigorous training and standardized protocols, on-the-field monitoring, and interim quality control of data, resulting in insignificant effects on results.

### Implications and recommendations for the humanitarian system

Working by the humanitarian principle of humanity entails striving to achieve equity across the different vulnerable segments that the response strives to serve. Lebanon’s “no-refugee policy” [20], compounded by deteriorating socioeconomic and political crises, exacerbates Syrian refugees precarious living conditions, under which older refugees immensely suffer. While significant efforts have been exerted by the government and the humanitarian eco-system to provide access to services to refugees in general, needs remain substantial and age-conscious interventions are lacking.

Two overarching considerations should be adopted by the humanitarian system to be more inclusive of older refugees. First, a paradigm shift must take place towards the more sustainable programming in the humanitarian-development nexus appropriate for a protracted crisis setting. This consideration resolves key issues affecting the lives of all residents, including the most vulnerable such as older refugees. Second, age-sensitivity must be mainstreamed as a cross-cutting consideration across all levels and phases of the humanitarian response. This starts with funding; since only less than 1% of humanitarian funding worldwide addressed elderly needs [16], humanitarian actors must advocate for funding tailored and inclusive programs targeting older refugees. In addition, baseline and needs assessments must collect age-disaggregated data to “see” the issues and vulnerabilities specific to older people. Programming must be based on inclusive needs assessments and a participatory human-centered approach. This could employ a “twin-track approach” ensuring the inclusion of OP into programs directed at the general population, and designing interventions tailored to the specific needs and issues of the elderly. Furthermore, monitoring and evaluation must incorporate age-disaggregated data for all performance indicators in blanket programs, and specific indicators for the tailored interventions targeting older people in emergencies. Participatory approaches should be adopted across the different activities related to accountability to affected populations, increasing the sense of participation and resilience, and leveraging on existing skills and experiences. All these phases and steps must be governed by detailed and nuanced age-inclusive standards adopted by humanitarian agencies into their policies, the implementation of which is diligently monitored [22].

Following these considerations, sector-specific recommendations can be proposed to enhance inclusion of OSRs guided by the ADCAP HISs. Because of Lebanon’s no-camp-policy and ambiguous regulations, it is crucial to find local solutions and designs more fitting for a protracted crisis and which abide to UNHCR shelter standards of protection [54, 55]. Research and development efforts engaging the government, UNHCR, academic institutions, and donors may serve as a working team to produce appropriate solutions. Shelter actors should also strive to relieve the burden of high rents by budgeting for rent support. WASH facilities must be within the residential units where possible. Alternatively, in ITSs with shared sanitation units, households with older people must be prioritized in being located closer to them and accommodations must be in place to assist those with limited functional status. Humanitarian health actors must act to increase appeals for chronic medications and hospitalizations, a challenge that may hinder service provision [27]. Alternatively, health cash vouchers that cover transportation, consultation, hospitalizations in non-contracted PHCCs, hospitals, and dispensaries and for non-supported and unavailable medications may be a plausible solution for elderly [56]. In addition, and in response to an overstretched healthcare system and staff, the government could consider special policies for integrating Syrian healthcare labor in remote areas where Lebanese systems are more fragile and inadequate to cover local needs [57]. As for nutrition and food needs, it is evident that the food coping strategies come as a result of prioritizing rent and electricity bills as well as medications, which are all spent from the cash assistance card, leaving little for food expenditure especially in households whose only source of income is the cash transfer. Because the cash for food assistance is only granted to registered refugees, other parties and humanitarian actors may need to provide and cover for non-registered refugees [58]. A holistic and complementary approach between sectors is needed to alleviate these burdens by addressing the high rents and the health needs, leaving the cash-for-food assistance primarily and adequately for food expenditure, especially when cash assistance is also granted to non-registered refugees through other organizations. The presence of elderly people with exacerbated vulnerabilities such as need for chronic medications and disability must be given significant importance in household vulnerability assessments used for the selection of granted households [58].

Other modalities include community-based interventions such as community kitchens [59], especially in closed ITSs providing a sustainable solution if also coupled with a business dimension beyond the philanthropic mandate [60]. This would be very appealing to the younger and more functionally capable older women, especially in light of their lifetime experience cooking homemade meals and the limited income sources under work restrictions. Other sustainable livelihoods opportunities include agriculture projects which could leverage on the legal possibility and refugees’ farming experience, especially that the vast majority of ITS residents came from rural areas in Syria [61, 62]. These initiatives can improve refugee-host relations, food security, nutritional status, and income through additional agricultural small businesses selling products prepared by refugees from the farm to the fork, including school food programs [62, 63].

On another note, further research is needed to support the evidence-base that (1) shows the vulnerabilities of older people in disasters and protracted crises, (2) fills gaps in understanding their needs and the ways to address them, (3) designs and evaluates the relevance, effectiveness, and impact of inclusive interventions, and (4) designs the age-conscious monitoring and evaluation tools to be used during and after implementation [15, 49, 64].

## Data Availability

Data cannot be shared publicly because of confidentiality and privacy concerns of study participants who are a particularly vulnerable population group with potential security and safety concerns. The American University of Beirut Institutional Review Board (IRB) approved the study, and the consent document that the IRB approved assured participants that their data would not be shared beyond the research team and as aggregated data in publications. The de-identified data for the study may be made available to investigators who contact AUB in accordance with institutional policies. Please note that AUB policies require AUB investigators to retain custody of research data, unless Non-Disclosure Agreements (NDA) have been signed prospectively with investigators/collaborators in other institutions. You can also contact AUB IRB office for any additional inquiries related to human subjects' data for research purposes.

## Acknowledgements

Authors would like to first express their sincere gratitude to all participants interviewed in this study. We would like to also acknowledge the Union of Relief and Development Associations for facilitating the access of the research team to the Syrian refugee beneficiaries residing in the informal tented settlements and hosted communities, and the field data collectors of the survey. A special thank you to Mr. Amir Abbass and Ms. Amale Kassir who assisted in reviewing the survey tools and Mr. Rida Mourad who assisted in supervising field work and transcription of the key informant interviews.

